# Clinical EEG slowing induced by electroconvulsive therapy is better described by increased frontal aperiodic activity

**DOI:** 10.1101/2022.04.15.22273811

**Authors:** Sydney E. Smith, Vincent Ma, Celene Gonzalez, Angela Chapman, David Printz, Bradley Voytek, Maryam Soltani

## Abstract

Electroconvulsive therapy (ECT) is one of the most efficacious interventions for treatment-resistant depression. Despite its efficacy, ECT’s neural mechanism of action remains unknown. Although ECT has been associated with “slowing” in the electroencephalogram (EEG), how this change relates to clinical improvement is unresolved. Until now, increases in slow-frequency power have been assumed to indicate increases in slow oscillations, without considering the contribution of aperiodic activity, a process with a different physiological mechanism. Here we show that aperiodic activity, indexed by the aperiodic exponent, increases with ECT treatment. This increase better explains EEG “slowing” when compared to power in oscillatory peaks in the delta (1-3 Hz) range, and is correlated to clinical improvement. In accordance with computational models of excitation-inhibition balance, these increases in aperiodic exponent are linked to increasing levels of inhibitory activity, indicating that ECT might ameliorate depressive symptoms by restoring healthy levels of inhibition in frontal cortices.

## Introduction

Since its introduction as a treatment for psychotic disorders more than 60 years ago, electroconvulsive therapy (ECT) has been widely used to treat affective illness. Indeed, ECT is the most efficacious treatment for Major Depressive Disorder (MDD), and it is often the preferred treatment for individuals who have failed to respond to numerous antidepressant trials. Similarly, ECT is the preferred treatment option for patients who are severely ill and require rapid relief of symptoms (e.g., due to active suicidality, psychotic depression or depression associated with loss of appetite and severe weight loss). One study showed that up to 80% of patients with treatment-resistant depression responded to ECT and many achieved full remission of their symptoms and were able to resume normal functioning^1^. A multisite collaborative study involving depressed patients referred for ECT (29.5% of the patients expressed suicidal thoughts or reported suicidal acts) showed that after one week of ECT, 38% of the patients reported a decrease in suicidal thoughts, after two weeks 61% reported a decrease, and at the end of the treatment course 81% reported a decrease^2^.

Given how well established, widely used, and clinically effective ECT is for the treatment of psychiatric conditions, it is surprising that its precise neural mechanism of action has not yet been identified. This is, however, not to say that no neural effects of ECT have been identified. Perhaps the most widely-replicated observation is that ECT is associated with a “slowing” of the electroencephalogram (EEG). Specifically, patients who receive ECT display increases in “slower”, or lower-frequency, delta (1-3 Hz) and theta (3-8 Hz) band power^3–8^. This “slowing” is observable both acutely, after any single treatment, and longitudinally over the course of a multi-session course of ECT^9^. Despite this, the neural mechanism behind EEG slowing remains unclear. In addition, how this increase in spectral power in the low frequency range is related to clinical outcome remains ambiguous^10^: several studies link EEG “slowing” to clinical ECT improvement^4,5,11–13^, some show the inverse relationship^14,15^, and others find no significant relationship^7,16–18^.

Although EEG “slowing” is one of the largest and most consistent effects in the ECT literature, it is not the only electrophysiological feature that has been associated with ECT treatment. For instance, decreases in alpha power^6^, decreases in beta and gamma power^18^, and increases in total spectral power (< 30 Hz)^4,7,16^ have all been observed with ECT. We hypothesize that these effects are actually the result of a change in a single, often overlooked EEG process: a change in aperiodic activity (Fig. 2). Specifically, we hypothesize that the plethora of EEG alterations observed in depression and in ECT treatment may be more parsimoniously explained by a change in aperiodic EEG activity. This interpretation may help disambiguate past observations of changes in spectral band power while simultaneously being better grounded by theoretical models of the physiological generators of aperiodic activity^19^.

To understand aperiodic activity, it is important to contrast it with the more well studied periodic—or oscillatory—activity. Methodologically, aperiodic and oscillatory activity are both reflected in the power spectrum of EEG data. While oscillatory power is concentrated at specific frequencies, neural power spectra also exhibit a broadband 1/f^*χ*^ scaling, where power is inversely proportional to frequency. The aperiodic exponent, *χ*, characterizes the slope of the power spectrum (Fig. 2A). An increase in the aperiodic exponent would cause a “steepening” rotation in the power spectrum, producing a large, consistent increase in low frequency power (Fig. 2B). This rotation could additionally produce apparent increases in total power, and/or decreases in alpha, beta, and gamma power depending on the spectral rotation frequency, like the fulcrum of a lever^20,21^. Changes in the aperiodic component have been linked to cognitive and perceptual states^21,22^, development^23^, aging^24^, anesthesia^25^, and disease states such as ADHD^26^ and schizophrenia^27^. Aperiodic activity has also been linked to the physiological effects of deep brain stimulation treatment for major depressive disorder^28^.

Importantly, if changes in the aperiodic exponent are not controlled for in analyses, changes in spectral band power can be misinterpreted as changes in oscillatory power, even where no true oscillation exists. Recent methods allow for explicitly separating oscillations and aperiodic activity^20,29^. Because neural oscillations likely have fundamentally different physiological origins than aperiodic activity^30–32^, conflating the two can lead to false conclusions about the neurobiological underpinnings of a particular electrophysiological change, which in turn can impact the development of more effective, targeted treatments. Thus far, analyses of EEG power spectra from ECT studies have not looked at aperiodic activity, but rather have measured power in the delta and theta ranges, under the assumption that power in those bands is equivalent to the existence of an oscillation in that frequency. Here, we investigate the hypothesis that ECT produces an increase, or steepening, in the aperiodic exponent in a population of 9 patients with MDD receiving a 12-session treatment of ECT. We analyze aperiodic and oscillatory EEG from frontal electrodes before and after approximately every fourth ECT session, repeating this procedure periodically throughout a full course of ECT. To test the aperiodic hypothesis, we compare changes in the aperiodic exponent to changes in canonical delta band power and delta oscillatory power as measured via spectral parameterization. We find that, acutely, increases in the aperiodic exponent better explain the observed increases in delta band power post-ECT, as opposed to true increases in delta oscillation power. Furthermore, we demonstrate that the aperiodic exponent continues to increase longitudinally through a course of ECT treatment and that the magnitude of this increase is related to clinical symptom improvement, an effect not seen in canonical delta band power. These results identify aperiodic activity as an important mechanism in ECT, especially in light of the relationship of aperiodic activity to excitation inhibition balance^32^ and the cortical inhibition theory of depression^33^.

## Results

### Clinical effects

The severity of depressive symptomatology as measured by the QIDS decreased over the course of a 12-session ECT treatment for all nine patients included in the study (p = 7.42 × 10^−12^), demonstrating the efficacy of ECT as a treatment of MDD (Fig. 1C).

**Fig. 1.**
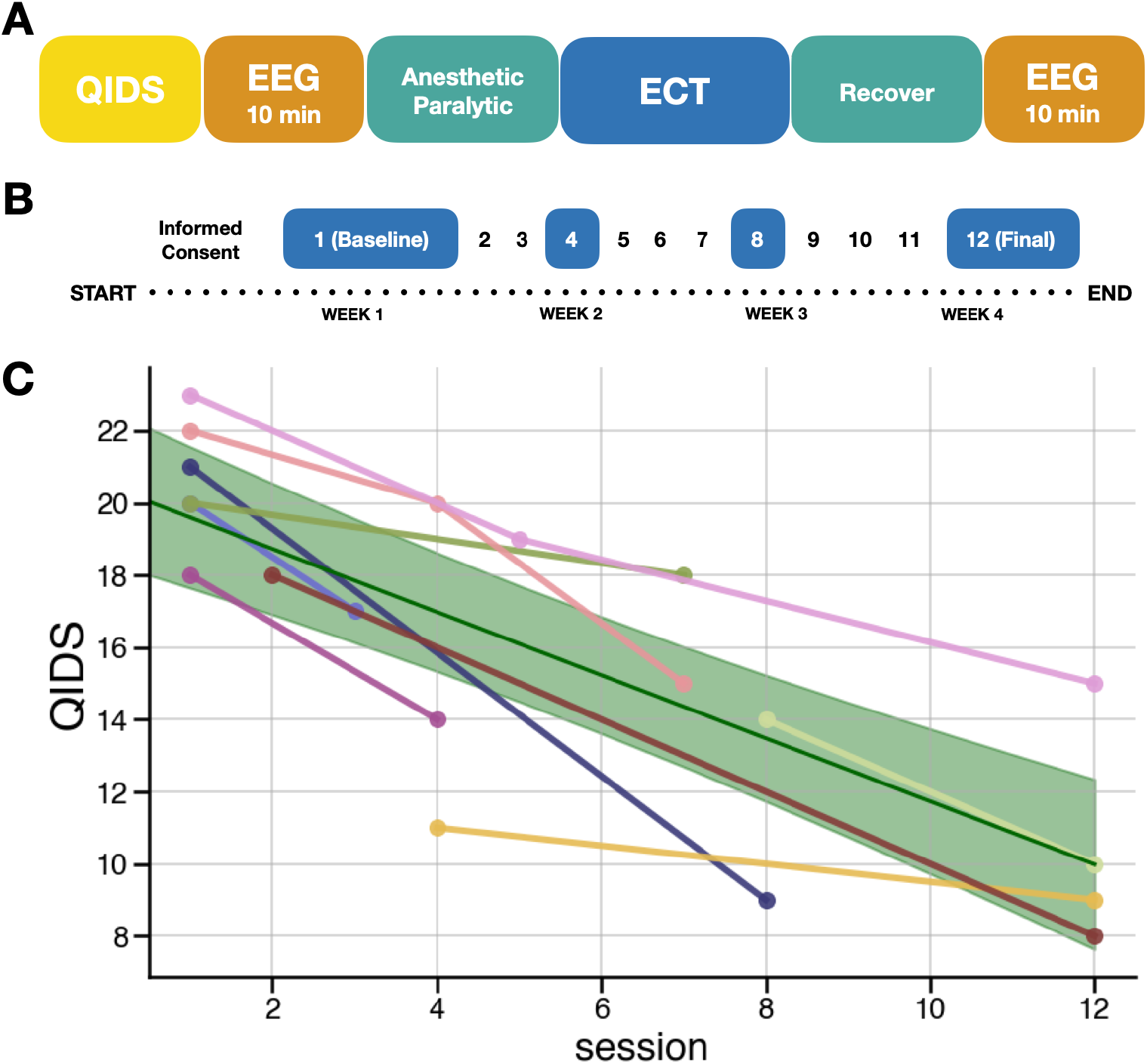
Schematic of the study. **a**, Single session process to capture acute effects of ECT on EEG. **b**, An example of the longitudinal protocol, in which the single session process for EEG is repeated at ECT treatment sessions 1, 4, 8, and 12. **c**, Clinical symptoms improve over the course of ECT treatment as measured by the Quick Inventory of Depressive Symptomatology (QIDS) scale (p = 7.42 × 10^−12^). Due to the constraints of recording in a clinical environment, a complete set of EEG recordings/clinical ratings was not possible for every subject, but subjects were included with a minimum of 2 sessions of pre and post ECT EEG recording.

**Fig. 2.**
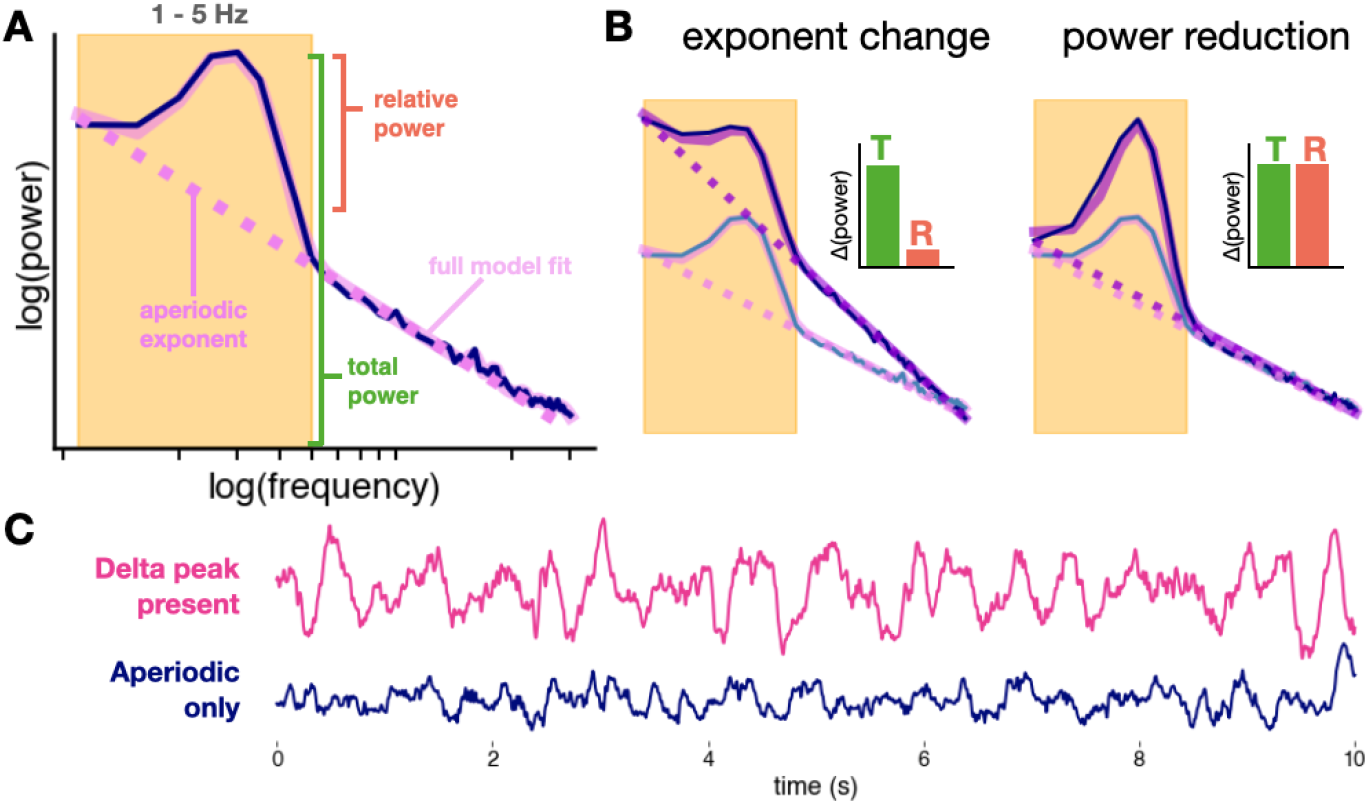
Hypothesis (aperiodic vs. oscillatory delta). **a**, Spectral parameterization quantifies the power spectrum as a composite of periodic and aperiodic components. Unlike traditional band power measures that conflate periodic and aperiodic activity, oscillatory power is defined as relative power above the aperiodic component (pink dashed line). **b**, Increases in the aperiodic exponent can cause apparent increases in total (T) band power, while power relative (R) to the aperiodic component remains unchanged. True increases in oscillatory power show increases in both total power and relative power. **c**, Delta in the EEG trace vs. aperiodic activity. EEG with delta oscillations (where a delta peak is present in the spectra) is visibly different from EEG with only aperiodic activity, though both can qualitatively look like EEG “slowing”. Only 4/9 EEG patients had delta peaks in their spectra for every session.

### Acute EEG effects

Following a session of ECT, EEG signals recorded from frontal electrodes displayed a significant increase in aperiodic exponent (d_z_ = 1.81, p = 9.20 × 10^−6^). This increase appears as a striking steepening of the power spectrum, with the average spectra across channels and subjects seeming to “rotate” around a frequency between 20-22 Hz (Fig. 3A-B).

**Fig. 3.**
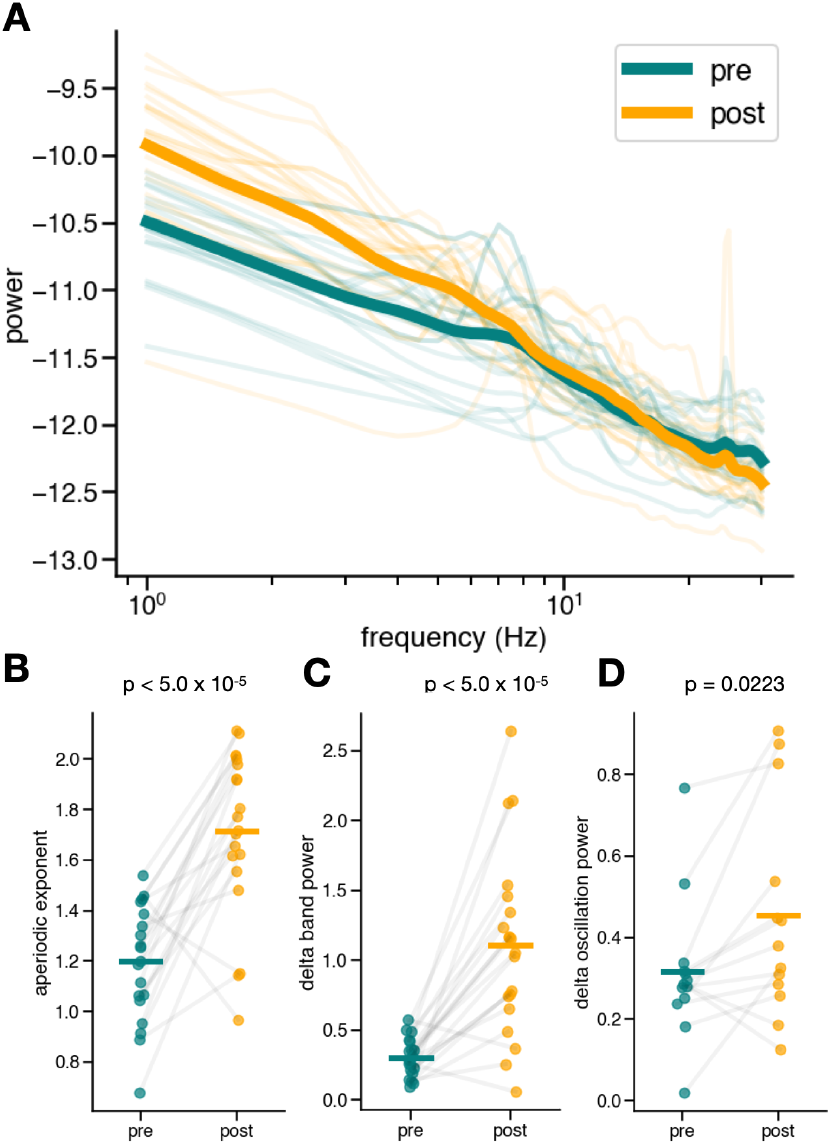
Acute effects. **a**, Power spectra of frontal electrodes pre- and post-ECT treatment. There is a visible “steepening” of the spectrum after ECT treatment. **b**, The aperiodic exponent of the power spectra pre- and post-ECT treatment, averaged across frontal electrodes for each patient. There is a large, highly significant (d_z_ = 1.81, p = 9.20✕10^−6^) increase in the aperiodic exponent for almost every patient. **c**, Delta (1-3 Hz) band power pre- and post-ECT treatment, averaged across frontal electrodes for each patient. When measured canonically, not controlling for the aperiodic exponent, a large, significant increase (d_z_ = 1.70, p = 1.59✕10^−5^) in delta band power appears. **d**, There is a small, yet significant increase in true oscillatory peak power in the delta range (d_z_ = 0.625, p = 0.0223). The effect size of delta band power increase is more similar in magnitude to that of the aperiodic increase than the effect size of the delta oscillation power.

We also observe the expected acute increase in delta band power (d_z_ = 1.70, p = 1.59 × 10^−5^) (Fig. 3C). However, the apparent EEG “slowing” effect is largely driven by the observed steepening of aperiodic activity; after controlling for changes in the aperiodic exponent (decomposing oscillatory and non-oscillatory estimates of PSD), oscillatory delta power changes were much less pronounced (d_z_ = 0.625, p = 0.0223) (Fig. 3D). The effect size of the increase in delta band power is comparable to that of the increase in aperiodic activity, more so than the effect size of the change in delta oscillation power. Crucially, only four out of the nine patients had a detectable delta oscillation in frontal electrodes for every session, highlighting the importance of spectral parameterization to detect oscillations. These results suggest that the well-replicated effect of ECT on delta band power is largely driven by the aperiodic exponent increase, instead of an increase in power of an oscillation in the delta band.

### Longitudinal EEG effects

The acute effect of ECT on EEG activity is potentially confounded by the effects of the anesthetic used during ECT treatment. Therefore, it is critical to also assess longitudinal changes of ECT treatment. To do this, we examined EEG activity across treatment days, before the introduction of anesthesia (Fig. 1). We found that the aperiodic exponent in frontal electrodes increases with repeated ECT treatments throughout a 12-session ECT course, as modeled by a linear mixed-effects model (p = 0.0178) (Fig. 4A-B). Unlike some previous studies that have found longitudinal increases in canonical delta band power, we find no significant effect of treatment on delta band power (p = 0.269) (Fig. 4D). This finding highlights the importance of spectral parameterization and its ability to disambiguate aperiodic and oscillatory activity, thereby allowing us to identify the longitudinal increase in aperiodic activity, a result which would have been impossible using only canonical band power.

**Fig. 4.**
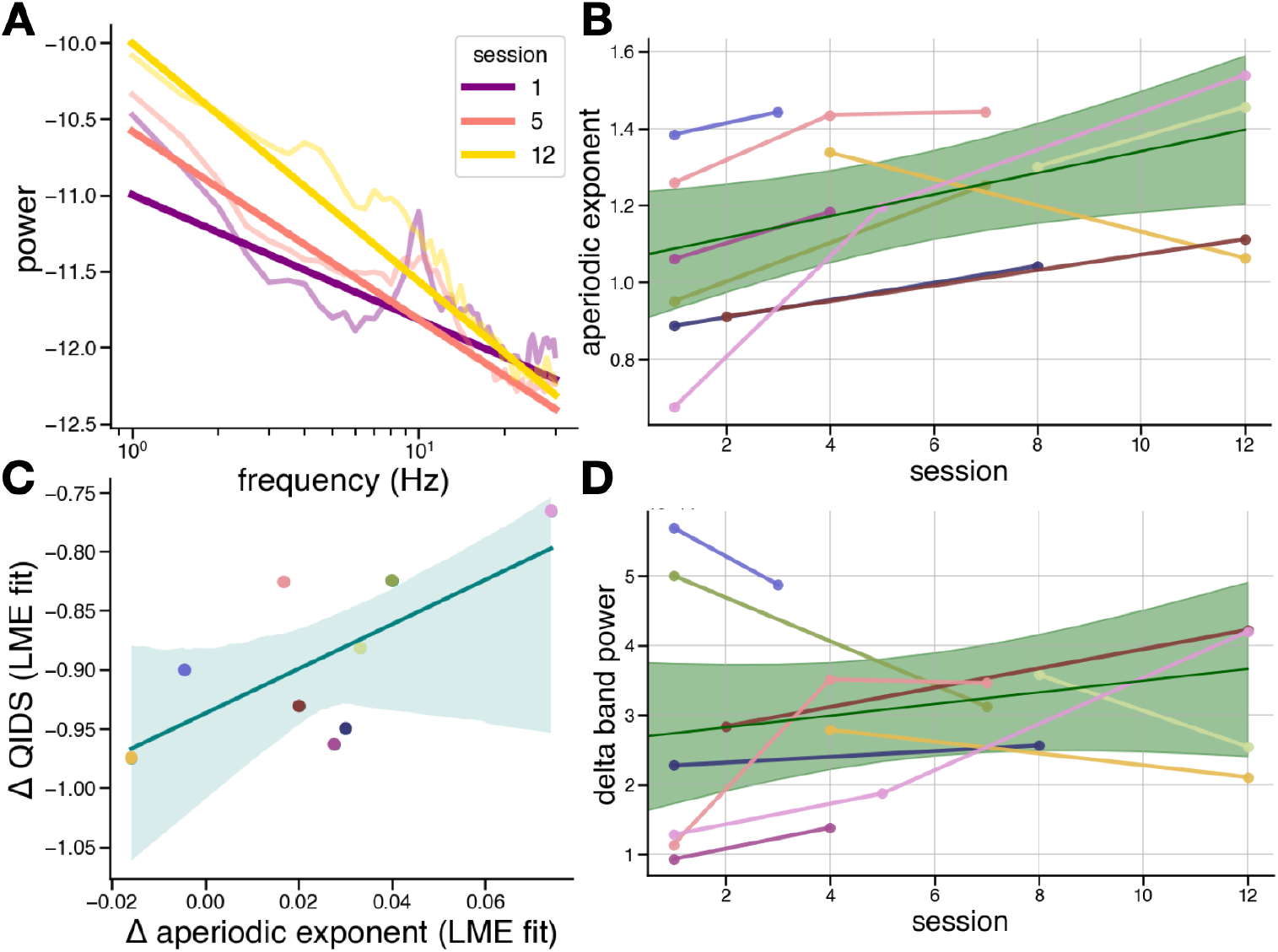
Longitudinal effects. **a**, Power spectra and aperiodic fits (straight lines) for average frontal PSDs for a single patient at three pre-treatment sessions. **b**, Aperiodic exponent significantly increases longitudinally (p = 0.0178). **c**, Across patients, improvements in QIDS scores, as quantified by the coefficients of a linear mixed-effects (LME) model fit to the data, are correlated to the increase in aperiodic exponent (r = 0.675, p = 0.0463). **d**, Delta band power does not significantly change longitudinally (p = 0.269).

### Clinical improvement and the aperiodic exponent vs. delta band power

The longitudinal increase in aperiodic exponent significantly tracks clinical improvement in this cohort of MDD patients receiving ECT, as indexed by QIDS assessment (Fig. 4C). We extracted the linear mixed-effects coefficients of each variable (aperiodic exponent and QIDS score) as it improved over repeated sessions for each patient. Each coefficient represents the rate of change for each variable with repeated treatments. We then performed a Pearson correlation of the rates of QIDS improvement and exponent increase for each patient and found a significant relationship (r = 0.675, p = 0.0463). Additionally, we found no significant correlation between clinical improvement and delta band power (r = 0.0856, p = 0.827). However, the limited size (n = 9) of the cohort included in this exploratory study should be considered in the interpretation of the results. More investigation is needed to definitively identify a relationship between changes in aperiodic activity and reduction of symptom severity for patients with MDD.

## Discussion

The observations that the aperiodic exponent increases as an acute effect of ECT and that it continues to increase throughout a course of multi-session ECT treatment connects long-standing EEG evidence to a potential mechanism of action. Although the exact physiological processes that produce the aperiodic activity indexed by the aperiodic exponent have not been precisely identified, one simple, yet promising, model suggests aperiodic activity in the local field potential (LFP) represents a summation of postsynaptic and transmembrane currents in a region^31^. A simplified model of excitatory glutamatergic and inhibitory GABAergic activity was able to capture changes in the aperiodic exponent^32^. In that model, the aperiodic exponent indexes the ratio of excitatory (E) to inhibitory (I) potentials present in the LFP, or excitation/inhibition (EI) balance. Specifically, the model indicates that a population of neurons with an EI balance biased toward a greater proportion of inhibitory activity would produce a more steeply sloped power spectrum and thus, a larger aperiodic exponent, than a population with an EI balance biased towards excitation. Normal changes in EI balance have been related to aspects of cognition, perception, and brain state^25^ and dysfunctions in EI balance have been linked to several neuropsychiatric and mood disorders, including MDD^34,35^.

This model of aperiodic activity and its relationship to EI balance is especially pertinent in the light of the cortical inhibition theory of depression. This theory states that patients experiencing symptoms of MDD have insufficient or dysfunctional inhibitory processes in various brain areas, including frontal cortices^33,36^. Specifically, these patients have insufficient GABAergic activity, reflected by reduced numbers of GABA neurons in the prefrontal cortex^37^. The prefrontal cortex plays an essential role in regulating EI balance in distributed networks throughout the brain^38^ and therefore, disruptions in EI balance in the prefrontal cortex could have wide-spread consequences affecting multiple networks, including limbic structures^39^ and the serotonergic and noradrenergic systems targeted by pharmacological antidepressants^40^.

The cortical inhibition theory has accumulated support from electrophysiology^41^, proton magnetic resonance spectroscopy^42,43^, and cell type-specific molecular genetics^44^, all of which highlight an abnormally low level of GABAergic activity in patients suffering from depression. Pharmacologically, this imbalance has been treated using factors that directly or indirectly reduce excitatory glutamatergic activity and increase inhibition with GABA agonists^34^. Treatments for depression, including ECT and serotonin-reuptake inhibitor antidepressants, have been shown to increase cortical GABA concentrations using proton magnetic resonance spectroscopy^45,46^. However, the link to how the electrophysiological changes seen with ECT could be reflecting changes in EI balance has not yet been illuminated^47^. In line with cortical inhibition theory, one proposed mechanism of ECT is that ECT ameliorates the symptoms of depression by directly inducing inhibitory activity over large portions of the cortex, causing widespread changes in functional connectivity and network dynamics^48^. The data presented here supports this proposed mechanism, suggesting ECT increases inhibition as indexed by the aperiodic exponent of the power spectrum.

Questions remain about whether the aperiodic exponent is a suitable non-invasive indicator for depression treatment response. Although we found a significant relationship between the degree of clinical symptom improvement and aperiodic exponent change, the directionality of the relationship indicates that larger increases in exponent are associated with smaller improvements in depressive symptomatology. We hypothesize that this result could indicate that patients who begin treatment with greater aperioridic activity might be more responsive to treatment. However, more research is required to investigate this relationship, especially because the size of the cohort included in this study was limited due to constraints of a clinical environment and restrictions around research during the COVID-19 pandemic, potentially leading to underpowered analyses of relationships between EEG and clinical improvement. Alternatively, at the relatively short timescales of a 4-week ECT treatment, ECT-induced restoration of inhibition might not be related to clinical improvement. For instance, a recent preprint using a method of estimating EI balance using spectral dynamic causal modeling for RS-fMRI found no relationship between treatment effectiveness and changes in EI balance when patients were scanned before and after treatment^49^.

Although the short timescale of this study prohibits us from drawing conclusions about long-term changes in aperiodic activity and its relationship to the clinical efficacy of ECT, other studies have shown abnormally elevated levels of slow frequency activity in EEG persisting weeks to months post treatment^16,50,51^. Like the results seen in this study, this increase in slow frequency power might be better explained by an increase in the aperiodic exponent, potentially suggesting a similar inhibition-related mechanism for ECT at longer timescales. Further investigation is needed to determine whether the aperiodic exponent could be used to predict treatment response from ECT as well as other treatment methods like TMS or various pharmacological interventions.

## Methods

### Participants

Nine patients with a diagnosis of MDD as per the Diagnostic and Statistical manual of Mental Disorders V were included in the study. Patients were recruited from the population of patients preparing to undergo ECT as part of their regular clinical treatment. Only patients with complete EEG recordings pre-and post-ECT treatment from at least two sessions were included in the present analysis. Written informed consent was provided by all patients and the study received ethical approval from the Veterans Association San Diego Health System (VASDHS) Institutional Research Board under the protocol H150012.

### Electroconvulsive therapy

ECT was administered three times per week according to VASDHS protocol. ECT was administered with a square wave, constant-current, brief-pulse device Thymatron System IV. Treatment was provided clinically at the discretion of the provider. Per standing clinical protocol, treatments were generally initiated at 5-10%, with subsequent treatments adjusted according to electrode placement (2x threshold for bilateral, 6x threshold for unilateral) with further adjustments as needed based upon seizure quality and duration (with a target of at least 30 seconds by EEG.) ECT was administered three times per week. See Table 1 for measures used per patient. Methohexital and succinylcholine were the typical anesthetic medications used (see Table 1). Treatment termination was based on response, clinical factors, and/or the patient’s expressed wish to discontinue ECT.

**Table 1.**
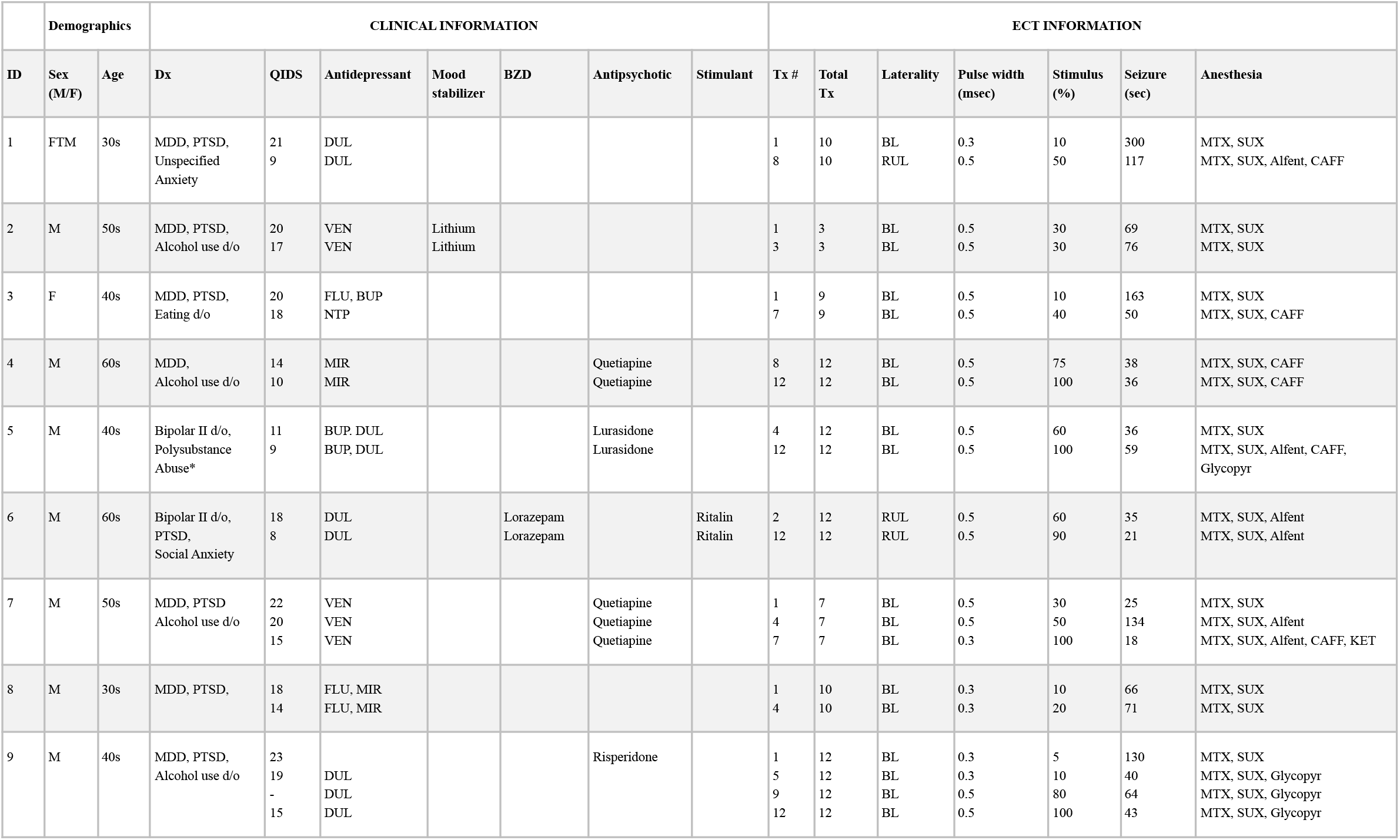
Patient demographics, clinical characteristics, and ECT treatment information. *Polysubstance abuse is no longer a DSM-V diagnosis. The patient for which that is listed was using Alcohol, Cannabis, Cocaine, Amphetamine and Opioid Dependence. All, apparently, in remission (not used for at least 1 year). *Abbreviations*: BUP (bupropion), ESC (escitalopram), DUL (duloxetine), FLU (fluoxetine), MIR (mirtazapine), NTP (nortriptyline), VEN (venlafaxine); MTX (methotrexate), SUX (succinylcholine), Alfent (alfentanil), CAFF (caffeine), Glycopyr (glycopyrrolate), KET (ketamine).

### Clinical measures

Quick Inventory of Depressive Symptomatology (QIDS)^52^ was administered by a clinician before each treatment session.

### EEG recording and data pre-processing

EEG recording procedures were designed to capture both acute effects of a single ECT treatment as well as longitudinal changes in EEG throughout the course of treatment (Fig. 1A). At the beginning of each session, 10 minutes of eyes closed, resting state EEG were recorded prior to administration of anesthetics or seizure induction. Following the seizure, patients were allowed to recover for at least 30 minutes until fully conscious and verbally communicative. This period was included in the study design in order to allow for recovery from the anesthetic medication as well as allow the research team to communicate with the patient about the recording procedure. This collection procedure was performed for multiple ECT sessions of the 4-week treatment (Fig. 1B). Due to constraints of the clinical environment, full recordings were not collected from every patient, but every patient included in the study had, at minimum, complete pre- and post-ECT recordings from two treatment sessions (Fig 1C). A 14-channel EEG headset device (Emotiv EPOC+, https://emotiv.com) containing saline-soaked felt pad sensors (online reference located at left/right mastoid) was used for all recordings. EEG data was sampled at 2048 Hz with internal downsampling at 256 Hz or 128 Hz and built-in notch filters at 50 Hz and 60 Hz for a bandwidth of 0.16 - 43 Hz.

EEG data was preprocessed to remove artifacts using a custom script in Python and MNE v.0.23.0^53^. First, channels with excessive noise and evidence of displacement were rejected manually based upon visual inspection. Data was then high-pass filtered at 1 Hz. To detect and remove ocular artifacts and eyeblinks, ICA was applied to the filtered data. Components containing ocular artifacts were removed. Due to the small number of channels, ICA could not always remove ocular and muscular artifacts, so as a final step, data segments were selected manually. Data was inspected visually and artifact-free segments were selected. Once selected, these segments were then detrended and concatenated. The first 100 seconds of this concatenated data was used for analysis. We employed this procedure due to the amount of noise in the data and small number of channels in the device. Because this concatenation technique produced high-amplitude, high-frequency artifacts, we restricted our analyses to a low frequency range (1-30 Hz) to avoid any contamination of the power spectra.

### Data analysis

Data analysis was performed with python using numpy v.1.18.1^54^, scipy v.1.6.2^55^, MNE v.0.23.0^53^, neurodsp^56^, and specparam^20^. Statistics were performed with python using statsmodels^57^ and with R^58^ to calculate linear mixed models using the lme4 library^59^. Only frontal channels AF3, AF4, F7, F8, F3, and F4 were included in the analysis. The code needed to reproduce the analysis and figures is provided here: (https://github.com/voytekresearch/smith_ect)

### Computing power spectral density (PSD)

PSDs were estimated using Welch’s method^60^ using 1.0 s Hamming windows with 0.5 s overlap. Custom functions for this can be found in NeuroDSP^56^, an open-source digital signal processing (DSP) toolbox for neural time series.

### Spectral methods

The spectral parameterization method and toolbox were used for calculation of aperiodic exponents as well as oscillatory power^20^. In this model, the power spectrum is composed of two electrophysiological components: aperiodic and periodic (oscillation) activity (Fig. 2A). This approach permits the disambiguation of contributions from oscillatory and aperiodic activity to the power spectrum. The aperiodic component of neural power spectra is described by the exponent and offset. Oscillatory activity is described by the center frequency, power, and bandwidth of identified peaks. If delta peaks were detected in the 1-3 Hz range, delta oscillatory power was defined as the power in the peak relative to the aperiodic component.

For methodological comparison, canonical delta band power was calculated as the mean of spectral power in the delta (1-3 Hz) frequency band.

Spectral parameterization and canonical band power evaluation were performed on the power spectrum of each channel (AF3, AF4, F3, F4, F7, and F8) for each recording session (pre and post). For each session, each EEG feature (aperiodic exponent, delta oscillatory power, and delta band power) was averaged across all six frontal channels. For acute effects analysis, EEG features from each pre-ECT session were compared to those from the corresponding post-session. For longitudinal analyses, only features from pre-ECT sessions were used to ensure that longitudinal changes in EEG features weren’t contaminated by effects of lingering anesthesia or postictal states unrelated to treatment efficacy.

### Statistical analysis

#### Clinical effects

Statistical evaluation of clinical effects was performed using a linear mixed effects model fit to the total QIDS clinical rating score. Patients were included as a random effect and ECT session number as a fixed effect.

#### Acute EEG effects

Effects of any given session of ECT on the aperiodic exponent, delta oscillatory power, and canonical delta band power were statistically evaluated using related-measures t-tests. Specifically, pre-ECT values were compared to post-ECT values for each patient, independent of session number.

#### Longitudinal EEG effects

Similar to the statistical evaluation of clinical effects, evaluation of longitudinal effects of repeated ECT sessions on the three EEG features (aperiodic exponent, delta oscillatory power, and canonical delta power) was performed using linear mixed effects models. A linear mixed effect model was fit to each EEG feature with patient as a random effect and EEG feature as a fixed effect. Only pre-ECT values were included in the model to avoid the potential confounding effects of the anesthetic on EEG activity.

#### Associating EEG features with clinical effectiveness

To determine if a significant relationship between the aperiodic exponent and clinical rating score was present, we used the lme4 library in R to extract the coefficients for each variable from the linear mixed-effects models. This approach produced three coefficients for each patient, corresponding to a rate of change for QIDS score, aperiodic exponent, and delta band power. We then used a Pearson correlation to assess the relationship of QIDS score x aperiodic exponent and QIDS x delta band power.

## Data Availability

Data produced in the present study are not currently available.

https://github.com/voytekresearch/smith_ect

## Acknowledgements

Support: NIH National Institute of General Medical Sciences grant R01GM134363-01 (to B.V.), UCSD Pace Grant (to M.S.), Majda Grant (to M.S.), Veterans Medical Research Foundation Pilot Grant (to M.S.)

Thanks: We thank Itay Hadas, Natalie Schworonkow, Ryan Hammond, Eena Kosik, Michael Preston, Quirine Van Engen, and Andrew Bender for their advice and feedback on the manuscript.

